# Evaluation of plasma biomarkers for causal association with peripheral artery disease

**DOI:** 10.1101/2023.05.05.23289560

**Authors:** Pranav Sharma, Michael G. Levin, Derek Klarin, Benjamin F. Voight, Philip S. Tsao, Scott M. Damrauer

## Abstract

**Background:** Hundreds of biomarkers for Peripheral artery disease (PAD) have been reported in the literature, however, the observational nature these studies limit robust causal inference due to the potential of reverse causality and confounding. We aimed to perform a systematic review of plasma biomarkers associated with PAD followed by Mendelian Randomization (MR) to systematically address residual confounding and better understand the causal pathophysiology of PAD. Combining a systematic review with MR facilitates cross-domain verification of observational and experimental results.

**Methods:** We performed a systematic literature review for terms related to PAD and/or biomarkers using Pubmed, Cochrane, and Embase, followed by manual review to extract biomarkers and their direction of effect. To evaluate evidence for causality, we employed Two-sample Mendelian randomization (MR). We developed genetic instruments for the biomarkers by mapping them to genome wide association studies (GWAS) of circulating biomolecules aggregated by the IEU Open GWAS and deCODE projects. We tested the association of the genetic instruments with PAD using summary statistics from a GWAS of 31,307 individuals with and 211,753 individuals without PAD in the VA Million Veteran Program. We employed Wald ratio or inverse-variance weighted MR; weighted median and weighted mode methods were applied as sensitivity analyses.

**Results:** We identified a total of 1,993 unique papers related to PAD and biomarkers using extant genetic instruments, and MeSH terms across PubMed, Embase, and Cochrane. After filtering and manual review, 170 unique papers remained, mentioning 204 unique biomarkers. Genetic instruments based on publicly available data were developed for 175 biomarkers. After accounting for multiple testing by controlling the false discovery rate (q < 0.05), 19/175 (10.9%) biomarkers had significant associations with PAD. Of the 19 significant associations, only 13/19 (58.3%) had concordant directions of effects with published reports. These 19 biomarkers represented broad categories including plasma lipid regulation (HDL-C, LPA, Triglycerides, APOA1, EPA, APOB, APOA5, and SHBG), coagulation-inflammatory response (CD36, IL6-sRa, VWF, IL18BP, and CD163), and endothelial damage/dysfunction (HLA-G, NPPA, VCAM-1, CDH5, MMP1, and INS).

**Conclusion:** This systematic review paired with Mendelian randomization elucidates biomarkers with genetic evidence for causality relevant to PAD, and highlights discrepancies between published reports and human genetic findings. Conventional studies have previously highlighted biomarkers that have correlation to PAD but have not emphasized the causal pathobiology of this disease. Expansion of genetic datasets to increase the power of these analyses will be crucial to further understand the causal role of plasma biomarkers and highlighting key biological pathways in PAD.

## INTRODUCTION

Observational studies, coupled with experiments in preclinical model systems have long sought to identify biomarkers associated with peripheral artery disease (PAD) and demonstrate their causal role in disease pathobiology.^1,2,3^ These approaches have identified hundreds of putative biomarkers with plausible biological mechanisms. However, observational studies remain at risk of residual confounding, which may lead to biased effect estimates or incorrect associations. For example, the development of PAD can simultaneously occur with the progression of other diseases such as coronary artery disease (CAD), strokes, and myocardial infarction (MI).^4,5.6^ Additionally, PAD has numerous complications such as critical limb ischemia (CLI), ulceration, and poor wound healing.^7^ These concurrent disease processes have the potential to change plasma biomarker profiles in observational studies, leading potentially to reverse causal associations, and increase the risk of confounding when trying to identify plasma biomarkers that are specifically responsible for the pathogenesis of PAD.^8,9^ Further, pre-clinical models focused on drug discovery lack the ability to differentiate casual biomarkers, are costly, and may lack external validity when trying to identify causal biomarkers.^10,11,12^

Genome-wide association studies (GWASs) have identified thousands of genetic loci associated with circulating biomarkers and cardiovascular pathologies, including risk-variants specific to PAD.^13,14,15^ Mendelian randomization (MR) uses genetic variation as a natural experiment to investigate the causal relationship between potentially modifiable risk factors and health outcomes in observational data.^16,17^ MR has been shown to be less susceptible to bias from residual confounding and reverse causality than traditional observational study design. MR has previously highlighted the role of major lipoproteins and subfractions in PAD.^18^ In this investigation, we use MR to evaluate the putative casual role of a broad set of plasma biomarkers in the pathophysiology of PAD.

We aimed to 1) systematically evaluate the literature to identify putative biomarkers of PAD and 2) leverage large-scale genetic datasets and Mendelian randomization to test the casual association of these biomarkers with PAD.

## METHODS

We conducted a systematic review based on the Preferred Reporting Items for Systematic Reviews and Meta-Analyses guidelines (http://www.prisma-statement.org/, **Figure 1**). We then developed genetic instruments for the putative biomarkers that were identified from the literature and tested their causal association with PAD using Mendelian randomization.

**Figure 1.**
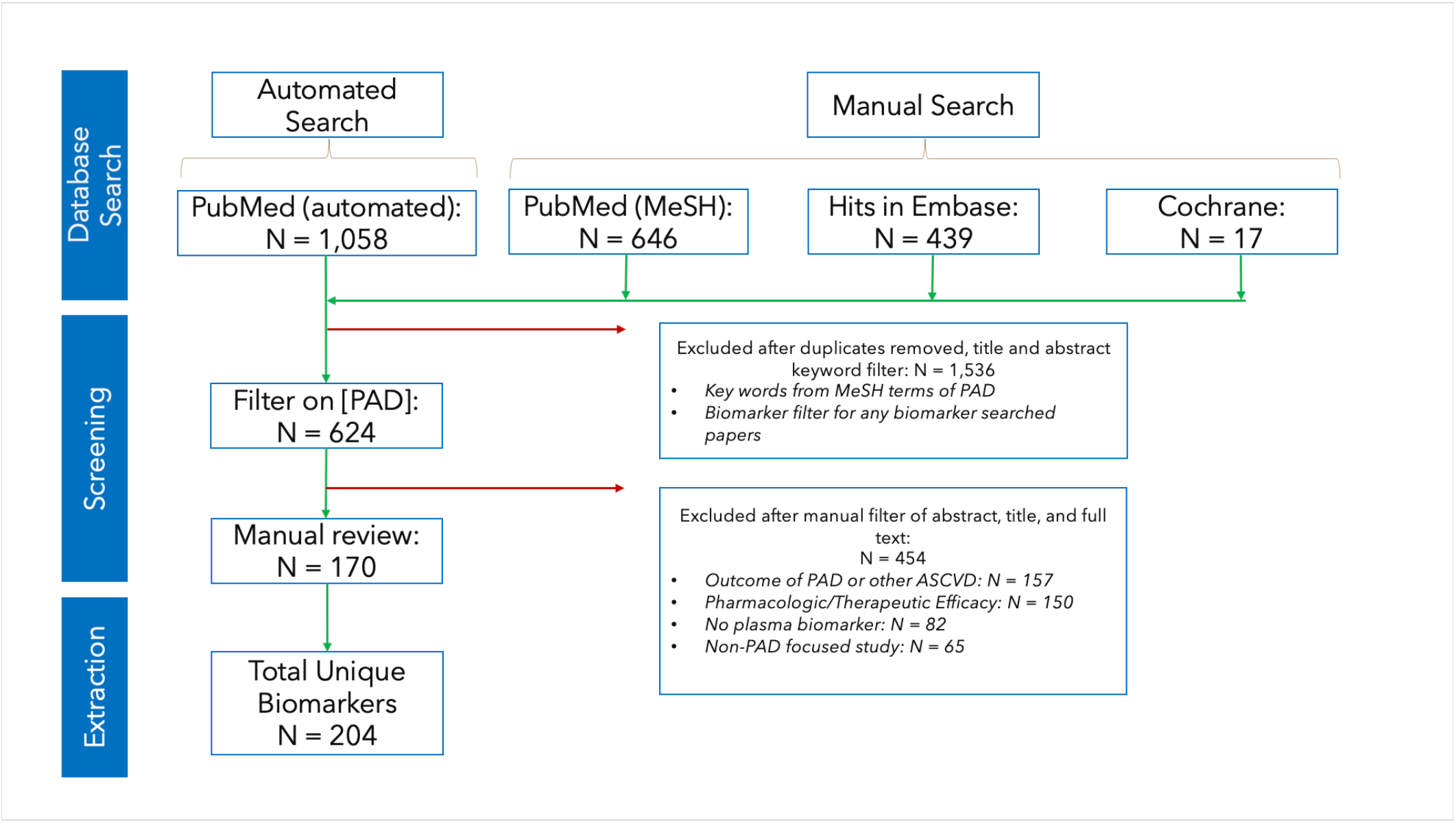
Automated and Manual Search and Selection process. N indicates the number of unique papers identified. After processing, the total number of unique plasma biomarkers identified were 204.

### Systematic Review of PAD Biomarkers

We conducted a systematic review of all literature referencing PAD and plasma biomarkers using two distinct methods. A systematic literature review was performed for MeSH terms of ‘PAD’ and ‘plasma biomarkers’ using PubMed, Cochrane, and Embase. All databases were searched in December 2021, and all relevant publications until this date were considered. A supplemental search was performed, focused on cardiovascular biomarkers for which there was extant genetic data to develop instruments for MR. This search method was based on the circulating protein GWAS cataloged on the IEU OpenGWAS project (https://gwas.mrcieu.ac.uk/) and the Olink cardiometabolic/cardiovascular protein biomarker lists (https://www.olink.com/products-services/target/). We leveraged the ‘pymed’ python package to query the PubMed API for ‘PAD’ and each biomarker found in the lists described above (https://pypi.org/project/pymed/). We included all study types that contained human and non-human samples and both prospective and retrospective analyses of individuals with PAD. Studies that reported the analysis of plasma biomarkers in the pathogenesis and progression of PAD were included. Studies that did not include PAD MeSH terms and keywords within titles and abstracts or observed non-PAD populations or analyzed drug effects on PAD populations were excluded. A detailed description of the approach including all key words is presented in **Supplemental Methods**.

### MR Genetic Instruments for Exposure data

Plasma biomarkers identified in the systematic review were mapped to genetic instruments found across a range of publicly available GWAS databases (Supplemental Table 1). For each biomarker, we selected genetic instruments from the largest available study to maximize sample size and power for subsequent Mendelian randomization. We constructed genetic instruments using independent genetic variants (r^2^<0.001, greater than 10,000 kb) that associated with the biomarker at genome-wide significance (P<5×10^−8^).

### MR Outcome data

Genetic instruments were tested for an association with PAD using summary data from the VA Million Veterans Program GWAS study (Klarin *et al*. 2019) which was comprised of 31,307 individuals with clinically diagnosed PAD (24,009 European; 7,373 African; and 1,925 Hispanic) and 211,753 individuals without clinically diagnosed PAD. This study defined PAD based on relevant diagnosis and procedure codes identified in the participants electronic health record.

Details of phenotyping, genotyping, and quality control have been previously described.^13^ The Million Veteran Program PAD genome-wide association study summary statistics were obtained from and available on dbGAP (Accession phs001672). There are no overlapping samples between MVP participants and individuals included in GWAS of the plasma proteome.

### Mendelian Randomization

Two-sample Mendelian randomization was performed using the *TwoSampleMR* (https://mrcieu.github.io/TwoSampleMR) package in R. The primary analysis used inverse-variance weighting with multiplicative random effects when multiple genetic instruments for a biomarker were available. When a single genetic instrument was available, Wald ratio MR was performed. Weighted median and weighted mode methods were applied as sensitivity analyses, as these methods make different assumptions about the presence of pleiotropy.^16,17^ To account for multiple testing we controlled the False Discovery Rate (FDR), defining significant associations with a threshold of q < 0.05. Additionally, the MR Steiger test of directionality was performed to validate direction of association between plasma biomarkers (exposures) and PAD (outcome).

### Gene Enrichment

ShinyGO (http://bioinformatics.sdstate.edu/go/) was used to perform gene ontology enrichment analysis on relevant plasma biomarkers. The comprehensive list of extracted biomarkers was mapped to gene symbols and an analysis highlighting the major pathways of observational and MR significant biomarkers was revealed through the analysis. The analysis was performed on a list of all biomarkers, all observational biomarkers excluding the significant plasma biomarkers from our MR analysis, and all significant biomarkers highlighted through our MR analysis.

### Bi-directional Mendelian Randomization

Elucidating the role of biomarkers in disease pathobiology can be difficult even with MR, and bi-directional relationships may be present.^19^ We repeated MR to test whether PAD could influence levels of these biomarkers. This method allowed us to evaluate if there was bi-directional association in between PAD and the biomarkers we identified as significant and helped refine the effect of PAD on plasma biomarker levels. We used the same exposure and outcome data to repeat the experiment, setting PAD (outcome) as exposure and significant plasma biomarkers (exposures) as outcomes.

## RESULTS

### Biomarker Identification

1,102 unique papers were found across PubMed, Embase, and Cochrane using MeSH terms to identify PAD biomarker studies. Secondary searches focusing on specific biomarkers for which genetic instruments could be derived identified an additional 1,058 articles. After removing duplicates and filtering on title and abstract inspection, 1,536 results were excluded. The remaining 624 articles were then reviewed for inclusion/exclusion criteria to identify studies that included plasma biomarkers that were relevant to the pathophysiology of PAD. After completing title/abstract review, the remaining 170 papers were examined further to extract a total of 204 unique plasma biomarkers and their direction of association to the development of PAD (**Figure 1, Supplemental Table 2**).

Of the 204 plasma biomarkers identified through our systematic review, 175 were mapped to unique genetic instruments and used as exposures (86%) (**Supplemental Table 3**). Circulating proteins were the predominant biomarkers identified. The remaining biomarkers included metabolites, such as circulating lipids, health measures, and cellular type ratios (i.e., Neutrophil Lymphocyte Ratio), found across publicly available GWAS studies. The number of variants associated with each plasma biomarker ranged from 1 to 408.

### Mendelian Randomization

Two-sample MR was used to estimate the effect of genetically predicted variation in each biomarker on the genetic liability to PAD (Supplementary Table 4). After accounting for multiple testing (FDR q < 0.05), 19 plasma biomarkers were significantly associated with risk of PAD (**Figure 2**). These 19 biomarkers represented broad categories including plasma lipid regulation (HDL-C, LPA, Triglycerides, APOA1, EPA, APOB, APOA5, and SHBG), coagulation-inflammatory response (CD36, IL6-sRa, VWF, IL18BP, and CD163), and endothelial damage/dysfunction (HLA-G, NPPA, VCAM-1, CDH5, MMP1, and INS). 13 of these had a direction of effect that was concordant with the published literature. The six biomarkers that did not show concordant direction of effect were: INS, EPA, NPPA, CD36, IL6-sRa, and CD163.

**Figure 2.**
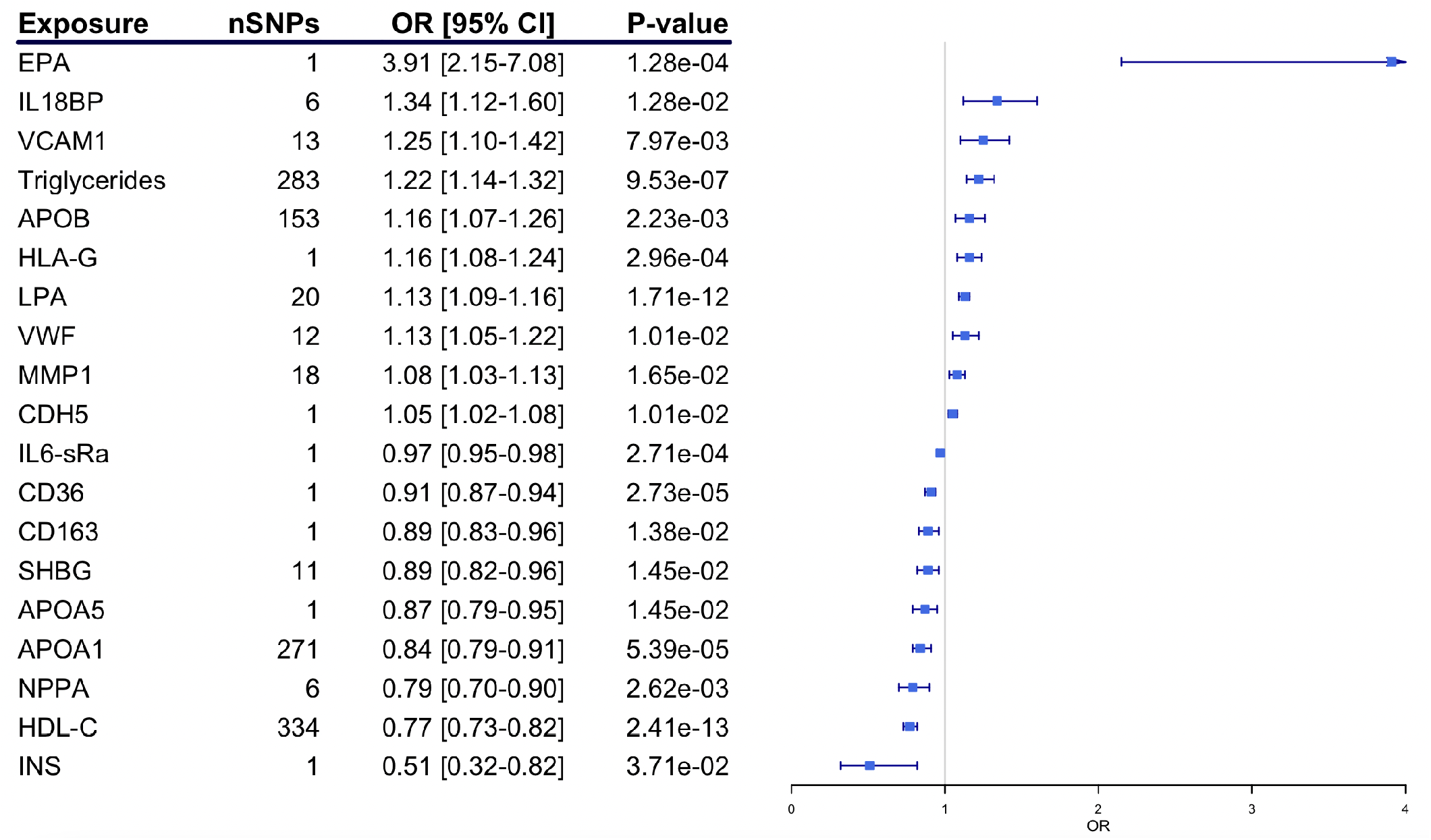
Mendelian Randomization Results highlighting significant plasma biomarkers. This analysis displays the 19 plasma biomarkers that highlight causal effect on the pathogenesis of PAD. Number of SNPs, Direction of effect, Standard error, FDR adjusted p-values, and OR with confidence intervals are shown for each biomarker.

Of the remaining 13 biomarkers, only 10 had enough variants in the instrument to perform sensitivity analyses. For these biomarkers, we used the weighted median and weighted mode methods as sensitivity analyses. All 10 had concordant effects and showed significance associations with PAD in at least one of these sensitivity analyses (p<0.05) (**Supplemental Table 5**).

### Gene Ontology Enrichment

To determine whether biomarkers identified in our systematic review and Mendelian randomization analyses were enriched for specific biological pathways, we performed gene enrichment analysis. An analysis on all biomarkers identified in our systematic review highlighted enrichment in multiple pathways including inflammatory response, negative regulation of multicellular organismal processes, and regulation of response to external stimuli (**Supplemental Table 6** and **Figure 3a**). Repeating this analysis, excluding all 19 significant MR plasma biomarkers, highlighted similar pathways such as inflammatory response, negative regulation of multicellular organismal processes, and cell activation (**Supplemental Table 7** and **Supplemental Figure 1**).

**Figure 3.**
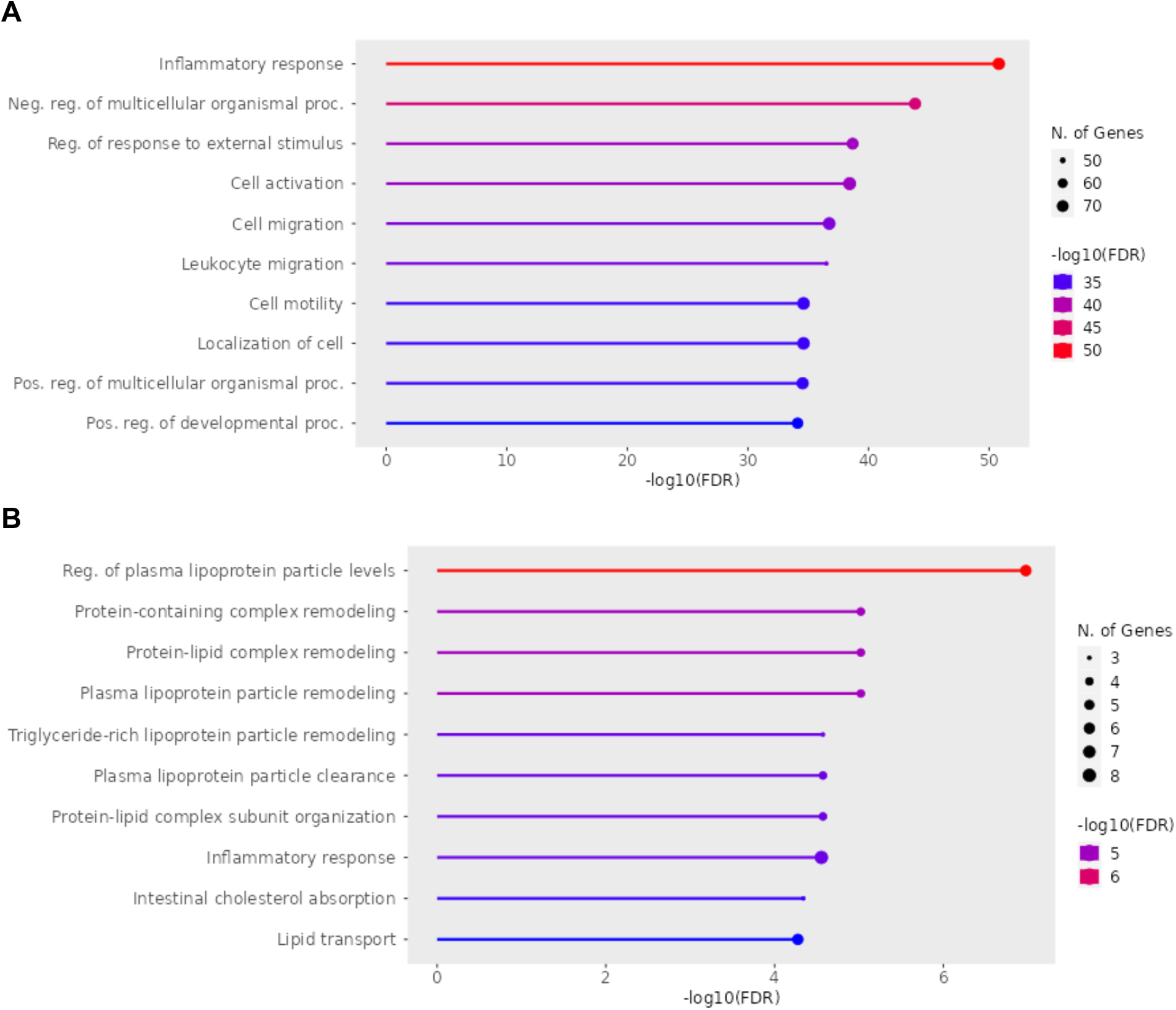
Gene Ontology Analysis of plasma biomarkers. The top 10 biological pathways prioritized by analysis of biomarkers identified from the observations literature (A) differ from those prioritized when considering the biomarkers for which there is causal evidence of association (B).

Biomarkers identified as significant in our MR analysis were enriched in a different set of biological pathways than those that were not, including pathways that involved lipoprotein metabolism, and inflammation (**Supplemental Table 8** and **Figure 3b**).

### Bi-directional Mendelian Randomization

To determine whether PAD might influence circulating levels of the biomarkers identified in our initial analysis, we performed bidirectional MR. This approach seeks to identify bi-directional relationships between exposures and outcomes (**Supplemental Table 9**). We found that 3 out of the 19 biomarkers identified in our analysis were influenced by genetic liability to PAD itself: MMP-1, IL18BP, and VCAM-1 (**Figure 4**). Given the bi-directional relationships for these 3 biomarkers, we utilized MR Steiger filtering to further verify the association of the biomarker as an exposure with the disease as an outcome and in all found support for genetic increases in the biomarker to lead to increased PAD risk (**Supplemental Table 10**).

**Figure 4.**
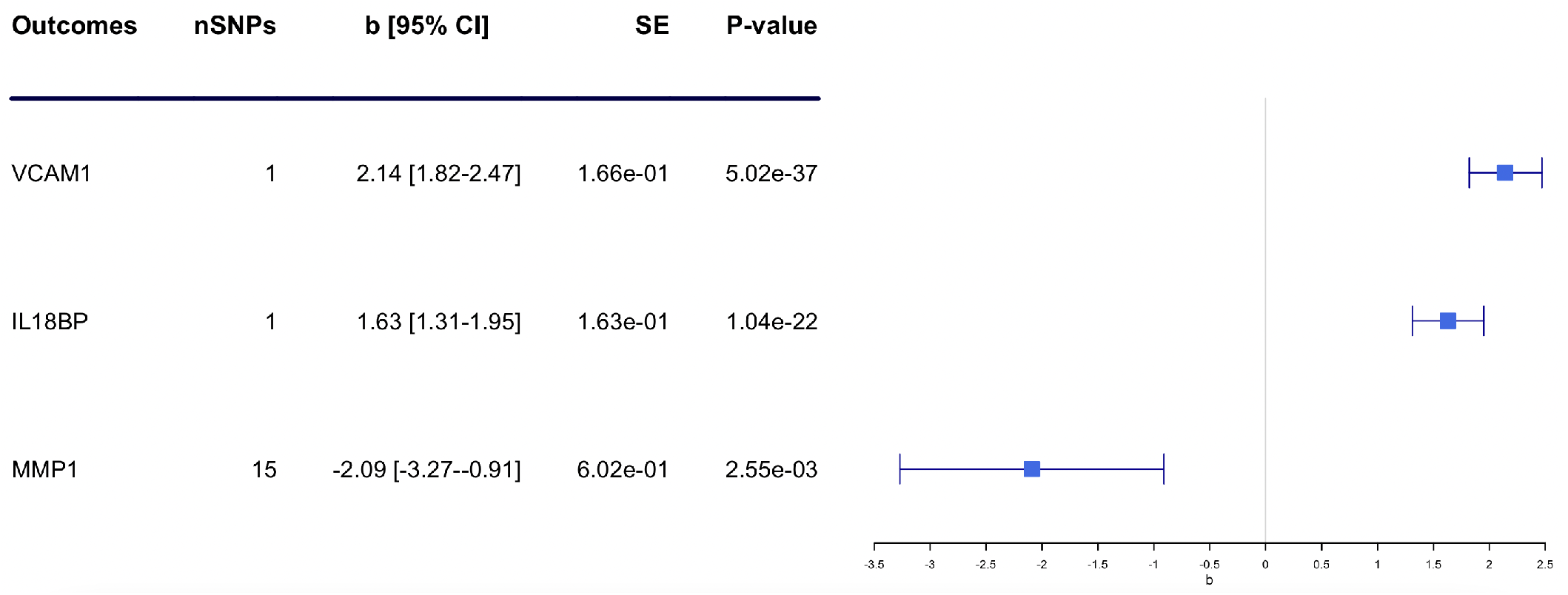
Bi-directional Mendelian Randomization Results highlighting significant bi-directional plasma biomarkers. This analysis displays the 3 plasma biomarkers that highlight a bi-directional effect in relation to PAD as an exposure and outcome. Number of SNPs, Direction of effect, Standard error, FDR adjusted p-values, and OR with confidence intervals are shown for each biomarker.

## DISCUSSION

This study aimed to provide a comprehensive overview of the role of plasma biomarkers in the pathophysiology of PAD. By drawing from several GWAS studies and building genetic instruments for identified plasma biomarkers, we were able to elucidate causal evidence for the association of circulating biomarkers to PAD. Overall, we found that although hundreds of circulating biomarkers have been associated in PAD by observational studies, only a small number have genetic support for a causal association.

Circulating biomarkers with genetic evidence for causal roles in PAD were concentrated in three central pathways: 1) plasma lipid regulation (HDL-C, LPA, Triglycerides, APOA1, EPA, APOB, APOA5, and SHBG), 2) coagulation-inflammatory response (CD36, IL6-sRa, VWF, IL18BP, and CD163), and 3) endothelial damage/dysfunction (HLA-G, NPPA, VCAM-1, CDH5, MMP1, and INS). These findings recapitulate known pathological pathways in the development of ASCVD and PAD.^20,21,22^

The biomarkers prioritized in our systematic review of the observational literature highlighted a different set of pathobiology as revealed by our gene ontology analysis. Of the biomarkers that were not significant in our MR analysis, the pathways of interest in observational literature were more broadly categorized into 1) inflammatory response, 2) multicellular response to external stimuli, and 3) cellular activation/immune response. One explanation for this discrepancy is that biomarker profiles observed in epidemiologic studies may be influenced by confounding factors reflected in common complications of PAD such as delayed wound healing, ulceration, and limb ischemia alongside other cardiovascular events.^23,24,25^

Even among those biomarkers for which there was both observational and causal support for their role in PAD, there were six exposures that did not report a concordant direction of effects between the MR analyses and the observed literature: CD163, IL6-sRa, EPA, CD36, NPPA, and INS. There are several possible mechanisms that can explain the discrepancy in reported results such as the endogenous processing of plasma biomarkers, lifetime exposure, or unique variants highlighting an unexpected effect compared to observational studies. For example, previous literature has explored the effects of exogenous EPA as a protective factor in the progression of ASCVD.^26^ Further, the REDUCE-IT trial reported that administration of EPA, 4g/d, resulted in fewer cardiovascular events (17.2% vs 22.0%; HR, 0.75 [95% CI, 0.68-0.83]).^27^ In contrast, our MR results suggest circulating EPA may increase risk. These MR findings might be highlighting a variant that leads to the improper processing of EPA, leading to higher endogenous plasma levels of this biomarker and inability to provide a protective effect, labeling EPA as a risk factor for PAD. Additionally, the relatively short-term effects assessed in traditional epidemiologic studies may in theory differ from those estimated by MR. Alternatively, these findings could suggest that there is a true positive direction of effect when considering plasma levels of EPA to PAD pathogenesis. Interestingly, there continues to be conflicting literature related to EPA supplementation benefits. More recently, the STRENGTH trial has shown that daily supplementation of omega-3 fatty acid formulation resulted in no significant difference in major adverse cardiovascular outcomes and was shown to increase tertiary end points (i.e., atrial fibrillation) and adverse gastrointestinal events when compared to corn oil-treated patients.^28^ Through this trial, they also reported that although omega-3 CA formulation is composed of both EPA and DHA, carboxylic acid formulation has greater bioavailability, permitting substantial elevations in EPA concentrations when compared to the REDUCE-IT trial, confirmed in phase 2 studies.^29,30^ It is unclear if the difference in increased levels of EPA in the blood, seen in these trials, has an impact on the risk of ASCVD. Methods such as mendelian randomization may provide more clarity into the causal effect of biomarkers on disease processes.

In sifting through observational studies and applying a new approach to differentiate casual biomarkers, we have shown the efficacy of Mendelian Randomization. The pathways highlighted through our gene ontology analysis, and more specifically some of the identified biomarkers, have been proven targets for successful therapy for ASCVD. For example, statins and PCSK9 inhibitors lower LDL-cholesterol, shown to increase risk of cardiovascular outcomes among individuals with PAD.^31^ Antithrombotic and anticoagulant agents like aspirin, clopidogrel, and rivaroxaban reduce major adverse limb events, and medications which act on the endothelium like cilostazol are used to improve symptoms. Several of the causal biomarkers we identified are specific targets of medications currently approved (ApoB, EPA), or in development (IL6-sRa, LPA and IL18B) for treatment of ASCVD.^32,33,34^ Previously successful pharmaceutical targets in this field have been validated in MR studies.^35,36,37,38^ These findings suggest the value of our approach when trying to understand the pathobiology of plasma biomarkers in their role of ASCVD, and more specifically, PAD.

Our studies identified 3 proteins with evidence of bi-directional associations with PAD: MMP-1, IL-18, and VCAM-1. MMP-1 is a matrix metalloproteinase secreted by many cells including fibroblasts, vascular smooth muscle and leukocytes, and has been shown to be upregulated in the breakdown of the extracellular matrix of tissue across the body.^39^ IL-18 is a pleiotropic cytokine involved in both innate and adaptive immune response and has been shown to be elevated in vascular pathologies such as CAD.^40^ Lastly, VCAM-1 is expressed within the luminal and lateral wall of endothelial cells during inflammatory processes and has been shown to promote monocyte chemotaxis in PAD.^41^ Interestingly, these are all cellular markers of inflammation.^42,43,44^ There are multiple clinical trials targeting the inflammatory effects of these biomarkers, particularly aimed at treated non-vascular diseases such as various cancers, Crohn’s disease, psoriasis, and rheumatoid arthritis.^45,46,47^ A majority of these trials are based on new clinical evidence highlighting the role of these biomarkers on particular disease processes, and the application of pre-existing drugs on therapeutic-slowing of disease progression. In terms of vascular disease, there are few studies looking at the therapeutic effects of inhibitors in this group of biomarkers.

With substantial investment in basic science and drug development, the translation to impact on patient care has been relatively uneven. A vast majority, ∼90%, of compounds that enter clinical trials fail to demonstrate sufficient safety and efficacy to gain regulatory approval.^48,49^ The failure of clinical trials in drug discovery is often due to the limited predictive value of preclinical models of disease. Mendelian randomization offers a solution to this issue by helping to uncover the causal role of biomarkers in the development of diseases like peripheral artery disease. Our study emphasizes the significance of exploring specific pathways in understanding the pathogenesis of PAD. MR avoids the limitations of observational studies by using the random allocation of genetic variants to determine causality. This method makes it possible to utilize existing drugs or discover new targets for the management and treatment of PAD.

### Limitations

This study should be analyzed within the context of its limitations. Firstly, the literature review has several limitations, mainly due to the heterogeneity of the underlying evidence. There was a lack of control across study-specific factors and the number of studies considered for biomarker discovery. Additionally, the study did not perform a meta-analysis of the literature and only captured individual-level study data.

Additionally, Mendelian Randomization has limitations of its own. MR makes assumptions about variants such as 1) association with lifetime exposure, 2) have independence from the outcome, and 3) only affect the outcome through the exposure and no other horizontally pleiotropic pathways.^50,51^ MR sensitivity analyses can test some of these assumptions, but the remaining assumptions are untestable. Beyond this, the effect estimates used to perform these analyses have the potential to be imprecise, leading to inappropriate magnitude of effect. Further, the conclusions of this study are limited by the availability and quality of genetic information, as well as the strength of the instruments used to measure the data. Lastly, the heterogeneity in the measurement of observational and genetic biomarker data makes it difficult to compare the magnitude of effect estimates.

Other limitations include certain constraints such as limited size and power of the utilized instruments. There is potential for residual confounding given the modest sample size of some significant biomarkers, which may account for incongruity in the direction of effects. However, as more biomarker datasets emerge, future studies can leverage increased power to refine these findings.

Instead, this technique should be taken as an estimate of casual association when exploring the effect of exposures on outcomes. Given these potential areas of limitation, this study should be interpreted as ongoing hypothesis generation related to the role of plasma biomarkers in the ongoing understanding of the pathogenesis of PAD and not be regarded as definitive evidence.

## Conclusion

This study highlights the importance of genetic techniques in elucidating the causal role of plasma biomarkers in the pathogenesis of PAD. The differences highlighted through gene ontology analysis emphasize the need to refine observational studies and explore genetic instruments to unveil key biological pathways in pathogenesis of PAD. As genetic datasets increase in size and availability, MR can be used to illuminate causal plasma biomarkers and pathways that should be explored further. Whether the biomarkers prioritized here represent novel therapeutic targets for PAD will require further functional validation.

## Data Availability

All data produced in the present study are available upon reasonable request to the authors

