## Supplementary material for "Evaluation of plasma biomarkers for causal association with peripheral artery disease": Supplementary_Materials_PAD_Final_Draft_SHARE.docx

**Systematic Evaluation of Plasma Biomarkers to Predict Their Causal Role in Peripheral Artery Disease Pathophysiology**

Scott M. Damrauer

Hospital of the University of Pennsylvania

Department of Surgery

3400 Spruce Street, Silverstein 4

Philadelphia, PA 19104 USA

**Table of Contents**

**Supplemental Tables (Supplemental Table File)**

**Supplemental Table 1.** Overview of Genetic Data Sets

**Supplemental Table 2.** Plasma Biomarkers and reported Direction of Effect

**Supplemental Table 3.** Genetic Instruments with Effect Size and Genome-Wide Significant SNPs

**Supplemental Table 4.** Inverse Variance Weighted and Wald Ratio MR Results for Plasma Biomarkers on PAD with FDR Adjustment

**Supplemental Table 5.** Total MR Results for Plasma Biomarkers on PAD

**Supplemental Table 6.** Gene Enrichment and Biological Pathways ShinyGO Analysis before MR

**Supplemental Table 7.** Gene Enrichment and Biological Pathways ShinyGO Analysis of Non-significant Plasma Biomarkers

**Supplemental Table 8.** Gene Enrichment and Biological Pathways ShinyGO Analysis of Significant Biomarkers after MR

**Supplemental Table 9.** Significant Biomarkers Results from Reverse MR analysis

**Supplemental Table 10.** MR Steiger test of Directionality

**Supplemental Table 11.** Total Harmonized Data

**Supplemental Table 12.** Total Harmonized Data (Bi-directional MR)

**Supplemental Methods**

**Supplemental Figures**

**Supplemental Figure 1.** Gene Ontology Analysis of all non-significant observational plasma biomarkers highlighted in MR analysis

**Supplemental Methods**

We conducted a wide search of all literature referencing PAD and plasma biomarkers using two distinct methods. First, a systematic literature review was performed for MeSH terms of ‘PAD’ and ‘plasma biomarkers’ using PubMed, Cochrane, and Embase. All databases were searched in December 2021, and all relevant publication until this date were considered. Second, we started our search with cardiovascular biomarkers for which there was extant genetic data to develop instruments for MR. This search method was based on the circulating protein GWAS cataloged on the IEU OpenGWAS project (<https://gwas.mrcieu.ac.uk/>) and the Olink cardiometabolic/cardiovascular protein biomarker lists (<https://www.olink.com/products-services/target/>). We leveraged the ‘pymed’ python package to query the PubMed API for PAD and each biomarker found in the lists described above (<https://pypi.org/project/pymed/>).

After collecting all papers from both search methods, the title and abstracts were filtered to narrow the analysis. The first search method was filtered for MeSH terms and keywords related to PAD on all titles and abstracts. The terms used included “Arterial Disease, Peripheral” or “Arterial Diseases, Peripheral” or “Disease, Peripheral Arterial” or “Diseases, Peripheral Arterial” or “Peripheral Arterial Diseases” or “Peripheral Artery Disease” or “Artery Disease, Peripheral” or “Artery Diseases, Peripheral” or “Disease, Peripheral Artery” or “Diseases, Peripheral Artery” or “Peripheral Artery Diseases” or “Peripheral Vascular Disease” or “Intermittent Claudication” or “Critical limb ischemia” or “Critical limb-threatening ischemia.”

The second search method was filtered for PAD MeSH terms, keywords, and the specific biomarkers used from the IEU OpenGWAS project and Olink cardiometabolic/cardiovascular protein biomarker lists on all titles and abstracts. The remaining studies were then screened by one author (P.S.), using the inclusion and exclusion criteria, defined below, for manual examination. Following this screen, relevant biomarkers were extracted, and duplicates removed.

After compiling all papers, and removing duplicates from these search methods, the title and abstracts were filtered to narrow the analysis. The first method was filtered on MeSH terms and keywords related to PAD on all titles and abstracts (Supplementary Methods/Table X).

The second search method was filtered for the same terms listed in Supplementary Table XX, and the biomarker of interest from the extant genetic data. For example, if the search contained the biomarker “Interleukin-6” and “PAD”, the results would be filtered on PAD terms and Interleukin-6. The remaining studies were then screened by one author (P.S.), using the inclusion and exclusions criteria defined below, for manual examination. Following this screen, relevant biomarkers were extracted, and duplicates removed.

We included all study types that contained human and non-human samples, both prospective and retrospective analyses of individuals with PAD. Studies that reported the analysis of plasma biomarkers in the pathogenesis of PAD were included. Studies that did not include PAD MeSH terms and keywords within titles and abstracts or observed non-PAD populations or analyzed drug effects on PAD populations were excluded.

**Supplemental Figures**

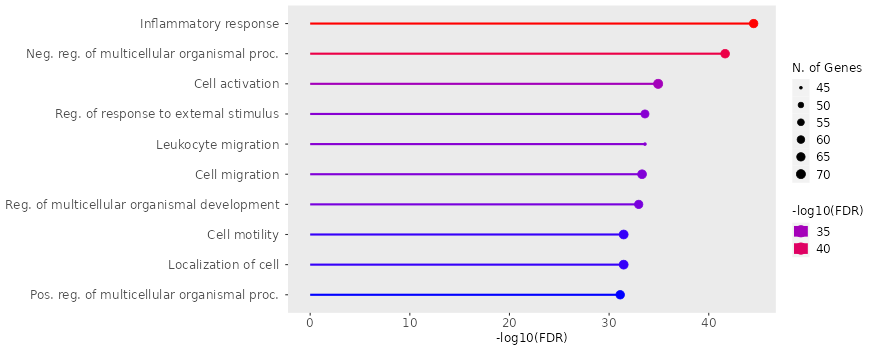

**sFigure 1. Gene Ontology Analysis of all non-significant observational plasma biomarkers highlighted in MR analysis.** This figure highlights the top 10 biological pathways of non-significant plasma biomarkers from our MR analysis.
